# Clinical Impact of Calcified Nodules in Patients with Severely Calcified Lesions Treated by Rotational Atherectomy: A Serial Optical Coherence Tomography Study

**DOI:** 10.1101/2023.12.29.23300649

**Authors:** Shuro Narui, Myong Hwa Yamamoto, Hiroyoshi Mori, Kisaki Amemiya, Toshitaka Okabe, Yui Koyanagi, Yuki Ito, Yuma Gibo, Takeshi Okura, Tatsuki Fujioka, Shigehiro Ishigaki, Soichiro Usumoto, Taro Kimura, Suguru Shimazu, Jumpei Saito, Yuji Oyama, Wataru Igawa, Morio Ono, Naoei Isomura, Masahiko Ochiai

**Affiliations:** Division of Cardiology, Showa University Northern Yokohama Hospital 35-1 Chigasaki-Chuo Tsuzuki, Yokohama, Kanagawa, Japan; Clinical Research Institute for Clinical Pharmacology and Therapeutics, Showa University 6-11-11 Kitakarasuyama Setagaya, Tokyo, Japan; Division of Cardiology, Showa University Fujigaoka Hospital 1-30, Fujigaoka Aoba, Yokohama, Kanagawa, Japan; Department of Pathology, National Cerebral and Cardiovascular Center 6-1, Kishibe-shinmachi Suita, Osaka, Japan

**Keywords:** percutaneous coronary intervention, calcified nodules, calcified lesions, optical coherence tomography, target lesion revascularization

## Abstract

**Background:** Percutaneous coronary intervention (PCI) for lesions with calcified nodules (CNs) is associated with worse outcomes than PCI for other calcified lesions. We aimed to clarify the relationship between CNs at index PCI, optical coherence tomography (OCT) findings at 8-month follow-up, and clinical outcomes using serial OCT.

**Methods:** This retrospective observational study utilized data from a prospective, single-center registry. We conducted consecutive PCI for calcified lesions requiring rotational atherectomy (RA) with OCT guidance. We categorized 51 patients (54 lesions) into those with (16 patients [16 lesions]) and without CNs (35 patients [38 lesions]).

**Results:** Post-PCI, stent expansion was comparable between the two groups, and CN-like protrusion was observed in 75% of patients in the CN group. Follow-up OCT at 8 months revealed in-stent CNs in 54% of treated CN lesions with CN-like protrusion, whereas non-CN lesions lacked in-stent CNs. The CN group exhibited greater maximum neointimal tissue (NIT) thickness than the non-CN group (p<0.001).

Multivariate linear regression analysis demonstrated that CN was associated with maximum NIT (p=0.02). Consequently, the CN group exhibited a higher clinically-driven target lesion revascularization (TLR) rate than the non-CN group at 1 (p=0.009) and 5 years (p=0.02). TLR primarily occurred in lesions with maximum CN angles >180°.

**Conclusions:** Following RA treatment with acceptable stent expansion, the presence of CNs before PCI correlated with greater neointimal tissue formation with in-stent CNs, resulting in a higher TLR rate, especially in lesions with maximum CN angles exceeding 180°.

*What is Known:*

- Calcified lesions with calcified nodules (CNs) are associated with a higher target lesion revascularization (TLR) rate in percutaneous coronary intervention (PCI) than other types of calcified lesions.
- In-stent CNs may appear after PCI for CNs and is a major cause of increased TLR rates after PCI for CNs.
- The relationship between calcified plaque morphology at the index PCI, neointimal tissue characteristics concerning in-stent CN location and frequency during follow-up, and subsequent clinical outcomes has not been explored with serial optical coherence tomography.

*What the Study Adds:*

- Follow-up OCT at 8 months revealed in-stent CNs in 54% of treated CN lesions with CN-like protrusion, whereas non-CN lesions lacked in-stent CNs.
- PCI for CNs exhibited a higher clinically-driven target lesion revascularization rate than PCI for lesions without CNs, primarily in lesions with maximum CN angles >180°.

## INTRODUCTION

Approximately 40% of patients undergoing percutaneous coronary intervention (PCI) exhibit moderate to severe coronary artery calcification, often requiring atherectomy, such as rotational atherectomy (RA), to facilitate stent delivery and expansion. However, calcified lesions still pose challenges due to increased risks of complications and adverse clinical outcomes, such as target lesion revascularization (TLR) and stent thrombosis.^1,2^

Among calcified lesions, eruptive calcified nodules (CNs), characterized by calcific nodules erupting in the lumen with disrupted fibrous cap,^3,4^ are associated with a higher TLR rate in PCI than other types of calcified lesions (those without CN).^5,6^ Compared with intravascular ultrasound, optical coherence tomography (OCT) provides superior imaging resolution, enabling precise evaluations of coronary calcification, particularly for CNs.^3^ Recent OCT studies have reported in-stent CN occurrences after PCI for CNs, confirmed by histopathological studies.^7,8^ Moreover, in-stent CN has emerged as a major contributor to the elevated TLR rate after PCI for CNs.^9,10^ However, the relationship between calcified plaque morphology at the index PCI, neointimal tissue (NIT) characteristics concerning in-stent CN location and frequency during follow-up, and subsequent clinical outcomes has not been thoroughly investigated using serial OCT examination.

This study investigated the relationship between CN occurrence at index PCI and in-stent CNs at 8-month OCT follow-up, the impact of CN occurrence at index PCI on clinical outcomes, and TLR-related CN characteristics.

## METHODS

### Study Design

This retrospective observational study originated from a prospective single-center registry at Showa University Northern Yokohama Hospital, Kanagawa, Japan. From April 2014 to August 2016, consecutive PCIs for severely calcified lesions requiring RA followed by stenting were performed under OCT guidance. Lesions with chronic total occlusion, myocardial infarction, coronary artery bypass grafting, or in-stent restenosis (ISR) were excluded (Figure S1). The institutional ethics committee of Showa University Northern Yokohama Hospital (Yokohama, Japan) approved the study protocol, and written informed consent was obtained from all patients. This study was conducted in accordance with the principles of the Declaration of Helsinki.

### PCI Procedures

An experienced operator (M.O.) conducted all rotational atherectomy (RA) procedures using a Rotablator (Boston Scientific, Maple Grove, Minnesota, MN, USA) under optical coherence tomography (OCT) guidance. RA was indicated by the following: (1) significant calcification evident through coronary angiography (moderate or severe), (2) inability to cross the OCT imaging catheter, or (3) severe calcium (Ca) findings on OCT (maximum angle of Ca ≥270°). All lesions were treated with RA, followed by pre-dilation, stenting using second-generation drug-eluting stent(s), and post-dilatation (when required). During the procedure, care was taken to avoid any decrease in rotational speed >5,000 rpm. After percutaneous coronary intervention, patients were administered dual antiplatelet therapy (aspirin and any P2Y12 receptor inhibitor) for 12 months, followed by single antiplatelet therapy.

### Angiographic Analysis

Angiographic analyses are described in Supplemental Material.

### OCT Imaging and Analysis

OCT images were acquired using the LUNAWAVE optical frequency domain imaging system and a FastView coronary catheter (Terumo Corporation, Tokyo, Japan). The OCT protocol encompassed pre-intervention (or pre-stent), post-PCI, and an 8-month follow-up after intracoronary nitroglycerin injection administration (0.2 mg). RA was performed before the initial OCT examination when the OCT catheter could not cross the lesion before a procedure. Qualitative and quantitative analyses were performed as previously described.^12,13^ The stented segment was considered as the lesion. Ca was defined as a signal-poor or heterogeneous region with sharply delineated borders. CN was defined as calcific nodules erupting in the lumen with fibrous cap disruption and substantive Ca proximal and/or distal to the lesion (Figure 1A-1).^3,4^ The maximum CN angle was assessed in the frame displaying the greatest CN angle. CN length was measured from the proximal to distal ends within the lesion. Malapposition was defined as a stent-adjacent vessel lumen distance >200 μm.^13^ Malapposition identified post-PCI but absent at follow-up was categorized as resolved; otherwise, it was considered persistent (Figure S2). A CN-like protrusion was defined as an irregular CN protrusion between stent struts, extending inside a circular arc connecting adjacent struts (Figure 1A-2) after stent implantation on the CN (Figure 1A-1). NIT was defined as at least five consecutive cross-sectional images with NIT exceeding 100 μm. An in-stent CN was defined as a new CN observed at the lumen side of the implanted stent, appearing as a high-backscattering protruding mass with an irregular surface, covered by signal-rich bands adjacent to the luminal surface over two consecutive cross-sections (Figure 1A–3).^9,14^ Maximum NIT thickness was defined as the thickest distance of the NIT within the lesion. The neointimal area percentage was defined as neointimal area/stent area × 100. All OCT findings were based on the consensus of the two observers (M.Y. and S.N.).

**Figure 1.**
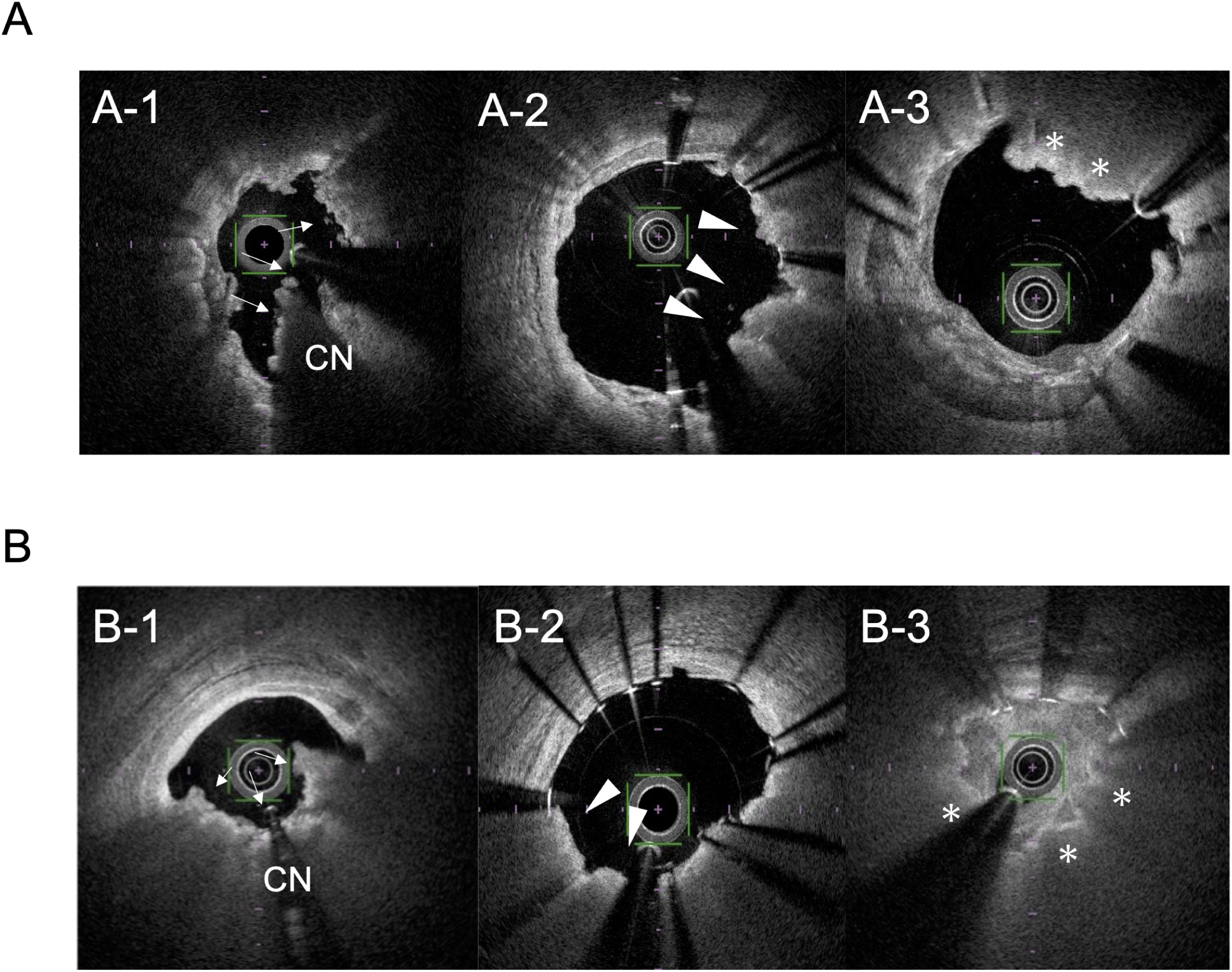
Representative OCT images. **(A)** Representative images of the OCT analysis definitions. CN is characterized by calcific nodules erupting in the lumen with fibrous cap disruption (arrows, CN) (A-1). CN-like protrusion is characterized by irregular CN protrusion between stent struts (arrowheads) (A-2) after stent implantation on the CN (A-1). In-stent CN is characterized by the appearance of a new CN on the lumen side of the implanted stent over two consecutive cross-sections (asterisks) (A-3). **(B)** A representative case of OCT images of a CN lesion modified by rotational atherectomy that required target lesion revascularization with an in-stent CN. CN lesion was modified using rotational atherectomy (arrows, CN) (B-1). Immediately after index PCI, acceptable stent expansion (79.6%) was obtained, and a CN-like protrusion was observed (arrowheads) (B-2). After 8 months of follow-up, an in-stent CN (asterisks) appeared with severe stenosis (B-3). CN, calcified nodule; OCT, optical coherence tomography.

**Figure 2.**
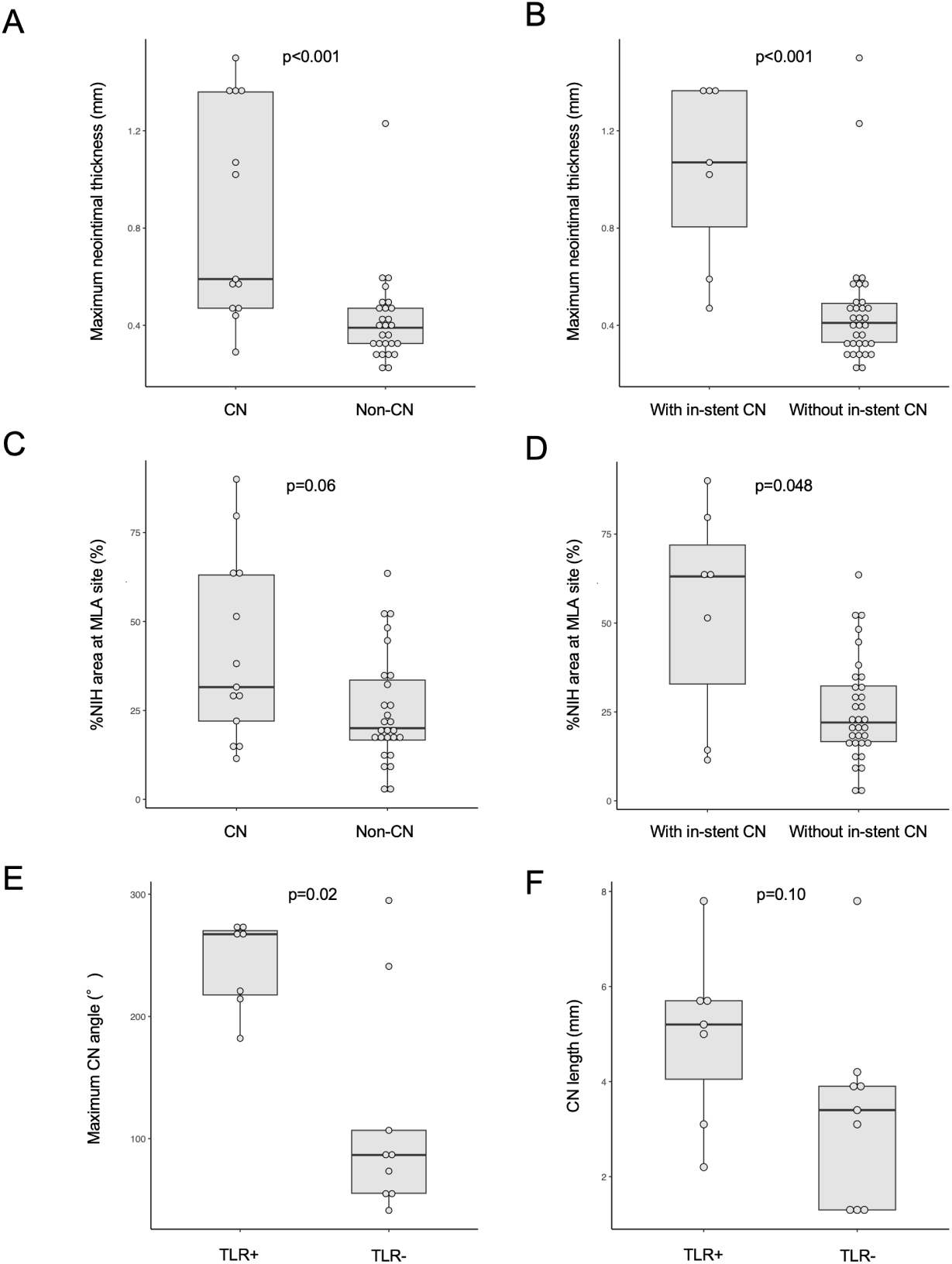
Difference in maximum neointimal thickness, % neointimal area at minimum lumen area site, and CN angle and length. Differences in maximum neointimal thickness between lesions with and without CNs (A) and between lesions with and without in-stent CNs (B). Difference in % neointimal area at the MLA site between lesions with and without CNs (C) and between lesions with and without in-stent CNs (D). Differences in the maximum CN angles between lesions with and without 5-year TLR (E) and differences in CN lengths between lesions with and without 5-year TLR (F). TLR, target lesion revascularization.

### Clinical Follow-Up

All patients were encouraged to receive a follow-up coronary angiogram and OCT 8 months after PCI regardless of their symptoms. They were followed up for 5 years through medical record review or communication with the patients or their families. Major adverse cardiac event (MACE) was a composite of cardiovascular death, myocardial infarction, and clinically-driven TLR. Cardiovascular death was defined as death caused by cardiovascular diseases, including myocardial infarction, sudden death, stroke, or heart failure.^15^ Myocardial infarction was defined according to the Fourth Universal definition.^16^ Clinically-driven TLR was defined as percutaneous or surgical revascularization recurring for a lesion within the stent or the 5-mm borders proximal or distal to the stent with at least one of the following: (1) fractional flow reserve <0.80, (2) diameter stenosis >50% (based on the average of multiple views) with recurrent symptoms or positive noninvasive functional test, or (3) diameter stenosis >70% (based on the average of multiple views), regardless of other criteria.^15^

### Statistical Analysis

Statistical analyses were conducted using JMP Pro 16 (SAS Institute, Cary, NC, USA) and R statistical software (version 4.2.1; R Foundation for Statistical Computing, Vienna, Austria). Continuous variables were presented as medians and first and third quartiles and compared using the Mann–Whitney U test. Categorical variables were expressed as frequency (%) and compared using the χ^2^ statistics or Fisher exact test. Multivariate linear regression models were used to adjust for clinically relevant baseline variables based on proven associations with large maximum NIT thickness. Long-term outcome estimates were assessed using the Kaplan–Meier method and compared using the log-rank test. The area under the receiver operating characteristic curve was estimated as the best cut-off value of the CN maximum arc at index PCI and as a TLR predictor. Statistical significance was set at p<0.05.

## RESULTS

### Study Population

Overall, 51 patients (54 lesions) were categorized into two groups: CN (16 patients and 16 lesions) and non-CN (35 patients and 38 lesions) (Figure S1). Follow-up OCT was obtained in 81% of CN (13/16) and 71% of non-CN lesions (27/38). The median duration from the index procedure to the follow-up OCT examinations was 7.8 (7.0– 9.4) months.

### Baseline Patient, Angiographic, and Procedural Characteristics

Patients in the CN group tended to be older (74 [71, 77] vs. 70 [59, 77] years, p=0.09) and had higher frequencies of renal insufficiency and hemodialysis than those in the non-CN group (Table 1). CNs were predominantly observed in the right coronary artery, whereas Ca lesions without CNs (non-CNs) were observed in the left anterior descending artery (Table S1). No significant differences were observed in angiographic and procedural characteristics between the two groups except for in-stent diameter stenosis at 8 months follow-up (21.0% [17.5, 51.6] vs. 17.0% [11.0, 23.0], p=0.04).

**Table 1.**
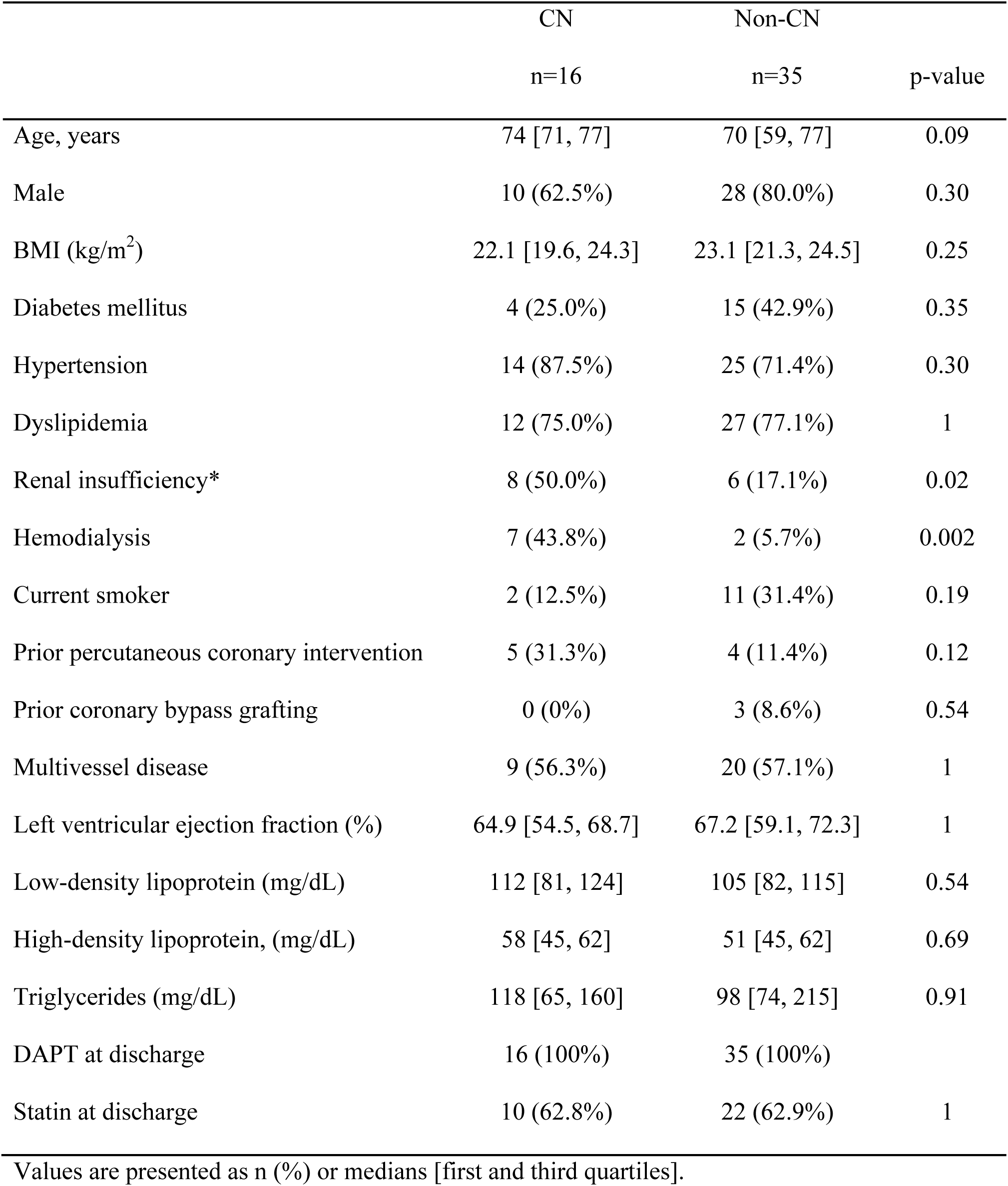
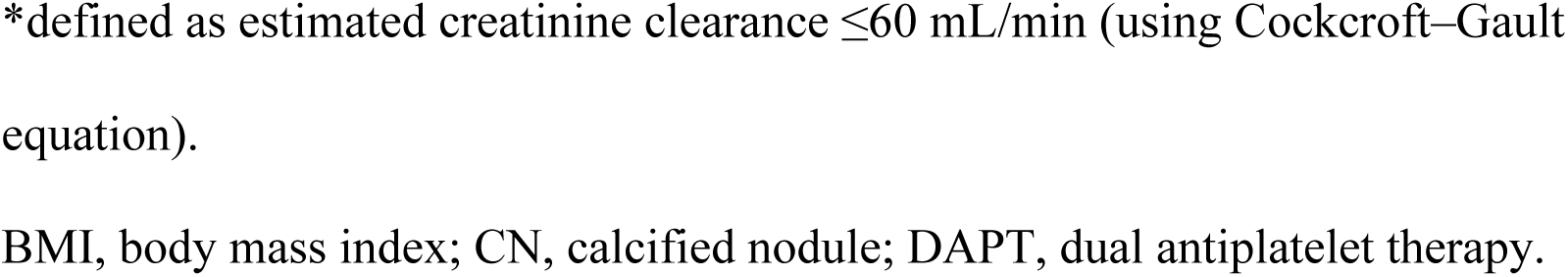
Patient characteristics.

|  | CN | Non-CN |  |
| --- | --- | --- | --- |
|  | n=16 | n=35 | p-value |
| Age, years | 74 [71, 77] | 70 [59, 77] | 0.09 |
| Male | 10 (62.5%) | 28 (80.0%) | 0.30 |
| BMI (kg/m<sup>2</sup>) | 22.1 [19.6, 24.3] | 23.1 [21.3, 24.5] | 0.25 |
| Diabetes mellitus | 4 (25.0%) | 15 (42.9%) | 0.35 |
| Hypertension | 14 (87.5%) | 25 (71.4%) | 0.30 |
| Dyslipidemia | 12 (75.0%) | 27 (77.1%) | 1 |
| Renal insufficiency* | 8 (50.0%) | 6 (17.1%) | 0.02 |
| Hemodialysis | 7 (43.8%) | 2 (5.7%) | 0.002 |
| Current smoker | 2 (12.5%) | 11 (31.4%) | 0.19 |
| Prior percutaneous coronary intervention | 5 (31.3%) | 4 (11.4%) | 0.12 |
| Prior coronary bypass grafting | 0 (0%) | 3 (8.6%) | 0.54 |
| Multivessel disease | 9 (56.3%) | 20 (57.1%) | 1 |
| Left ventricular ejection fraction (%) | 64.9 [54.5, 68.7] | 67.2 [59.1, 72.3] | 1 |
| Low-density lipoprotein (mg/dL) | 112 [81, 124] | 105 [82, 115] | 0.54 |
| High-density lipoprotein, (mg/dL) | 58 [45, 62] | 51 [45, 62] | 0.69 |
| Triglycerides (mg/dL) | 118 [65, 160] | 98 [74, 215] | 0.91 |
| DAPT at discharge | 16 (100%) | 35 (100%) |  |
| Statin at discharge | 10 (62.8%) | 22 (62.9%) | 1 |

Values are presented as n (%) or medians [first and third quartiles].
\*defined as estimated creatinine clearance $\leq 60$ mL/min (using Cockcroft–Gault equation).
BMI, body mass index; CN, calcified nodule; DAPT, dual antiplatelet therapy.

### OCT Findings

The CN group more frequently required RA before OCT assessment than the non-CN group (11 [68.8%] vs. 10 [26.3%], p=0.006) (Table 2). The CN group had a longer Ca length, higher maximum Ca angle, and greater Ca thickness than the non-CN group. Post-PCI OCT findings indicated no difference in minimum stent area and expansion between the groups. CN-like protrusion was observed in 12 lesions (75%) within the CN group. In-stent CNs were observed in 7 lesions in the CN group but were absent in the non-CN group (53.9% vs. 0%, p<0.001). In the available cases with follow-up OCT, all in-stent CNs were located at sites where CN-like protrusion was observed in post-PCI OCT. The remaining four cases with CN-like protrusion transformed to homogeneous high-intensity NIT during follow-up (Figure S3). The CN group had a greater maximum NIT than the non-CN group (0.59 [0.47, 1.34] vs. 0.39 mm [0.32, 0.48], p<0.001). The CN group exhibited a larger neointimal area at the minimum lumen area (MLA) site than the non-CN group (3.2 [1.5, 4.5] vs. 1.3 mm^2^ [0.9, 1.7], p=0.007). The MLA site during follow-up coincided with the baseline CN location in 84.6% of the CN group. Moreover, the difference in maximum NIT and % neointimal area at the MLA site was more pronounced when comparing lesions with and without in-stent CNs (1.07 [0.59, 1.37] vs. 0.41 mm [0.33, 0.50], p<0.001; 63.1% [14.3, 79.7] vs. 22.0% [16.7, 33.5], p=0.048, respectively) (Figures 2A–D). Multivariate linear regression analysis revealed that CNs at index PCI were associated with a larger maximum NIT (regression coefficient, 0.303; 95% confidence interval, 0.057–0.549; p=0.02) (Table 3). While the frequency of malapposition at post-PCI OCT was comparable between the two groups, OCT at follow-up showed a trend toward a higher frequency of persistent malapposition in the CN group than in the non-CN group (Figure S2). Figure 1B shows a representative case illustrating OCT images of a CN lesion modified by RA that had CN-like protrusion just after PCI, requiring a subsequent TLR owing to an in-stent CN.

**Table 2.**
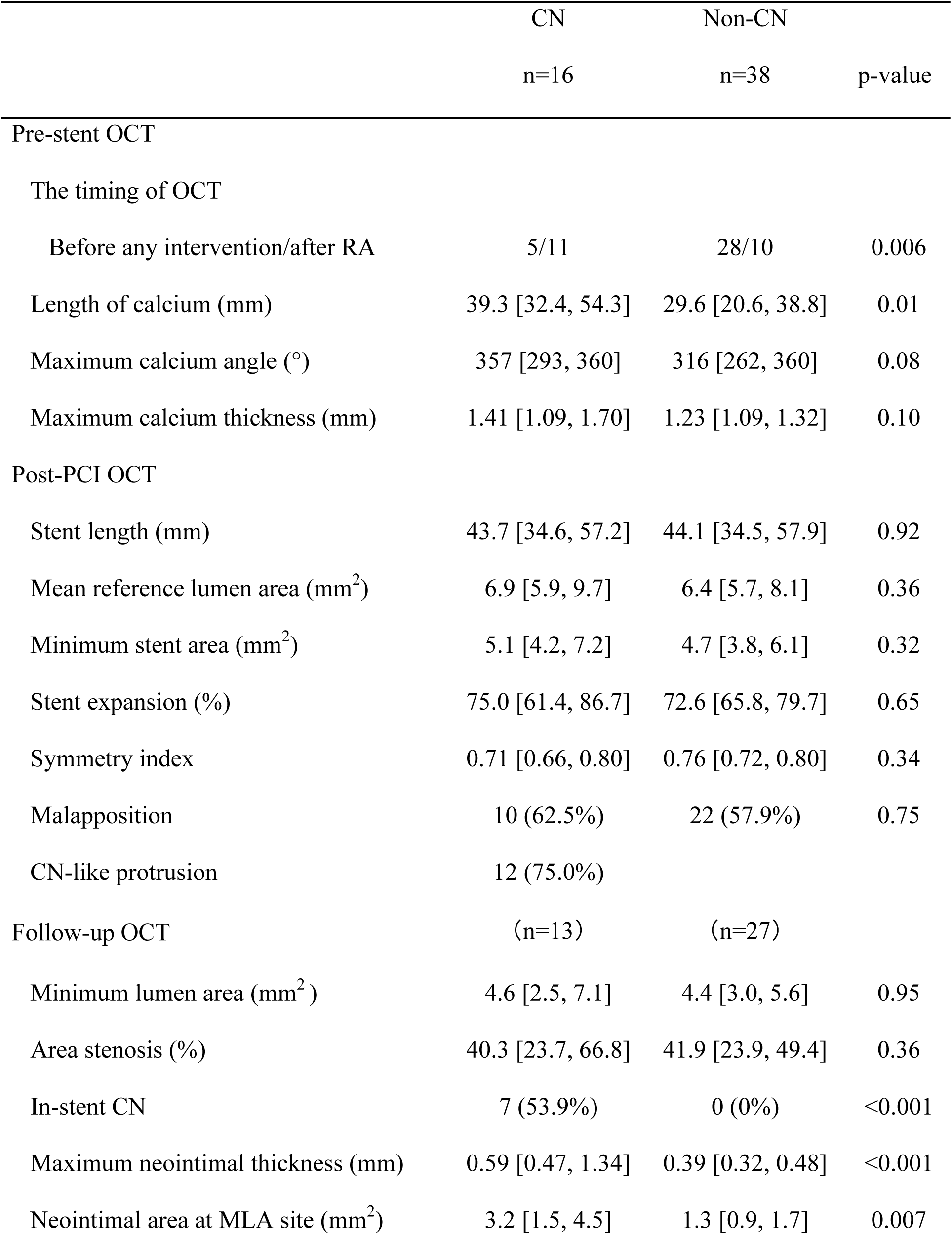
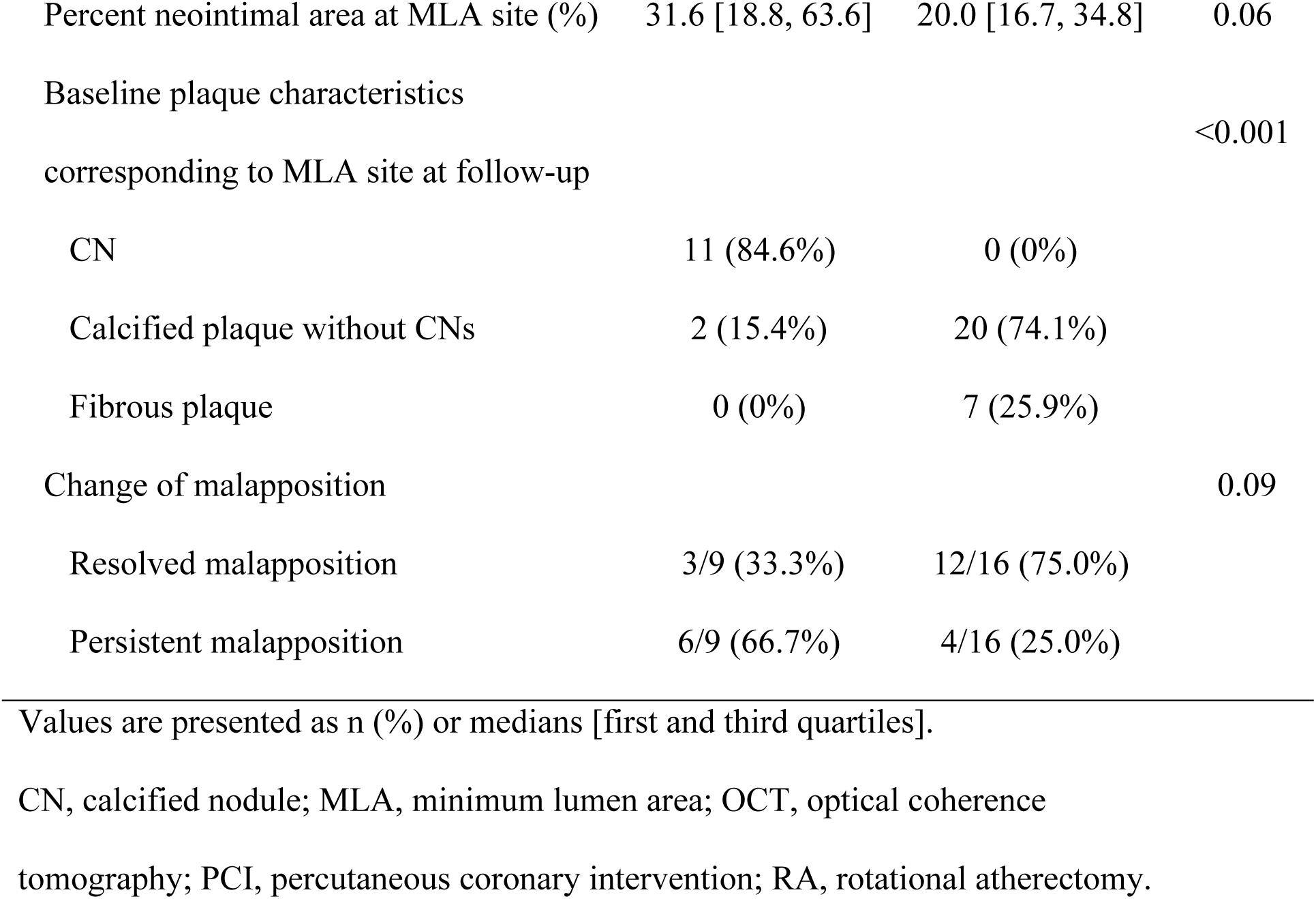
OCT findings.

**Table 3.** Multivariate linear regression analysis to predict large maximum.

|  | Multivariate |  |  |
| --- | --- | --- | --- |
|  | Regression<br>coefficient | 95% confidence interval | p-value |
| Age | -0.003 | -0.015–0.008 | 0.54 |
| Hemodialysis | 0.186 | -0.486–1.080 | 0.45 |
| Calcium length | 0.007 | -0.001–0.014 | 0.07 |
| Stent expansion | 0.004 | -0.005–0.012 | 0.36 |
| CNs at index PCI | 0.303 | 0.057–0.549 | 0.02 |

### Clinical Outcomes

The median follow-up period was 5.0 (1.7–7.0) years. The CN group experienced MACE more frequently than the non-CN group. The CN group had a higher TLR rate than the non-CN group at 1 year (31.3% vs. 2.9%, p=0.009) and 5 years (43.8% vs. 11.4%, p=0.02) (Table S2, Figure 3A). The median TLR in the CN group was 238 (151, 1112) days. The incidence of cardiovascular death at 5 years in the CN group tended to be higher than that in the non-CN group (25.0% vs. 5.7%, p=0.07). We further compared lesions with (n=7) and without 5-year TLR (n=9) in the CN group to clarify TLR-related CN characteristics at the index PCI. Lesions with 5-year TLR exhibited a greater maximum CN angle than those without (267° [214, 273] vs. 87° [55, 174], p=0.02) (Figure 3E). However, the CN length (5.2 [3.1, 5.7] vs. 3.4 mm [1.3, 4.1], p=0.10) (Figure 3F), maximum Ca angle (353° [279, 360] vs. 360° [317, 360], p=0.27), and Ca length (38.0 [35.0, 55.2] vs. 40.6 mm [23.0, 52.3], p=0.43) were comparable. Receiver operating characteristic curve analysis identified the optimal cut-off value for the maximum CN arc to predict 5-year TLR as 182° (sensitivity, 100%; specificity, 77.8%; p=0.003), with an area under the curve of 0.84. Figure 3B illustrates TLR-free survival, comparing CN angles ≥180° and <180° within the CN group.

**Figure 3.**
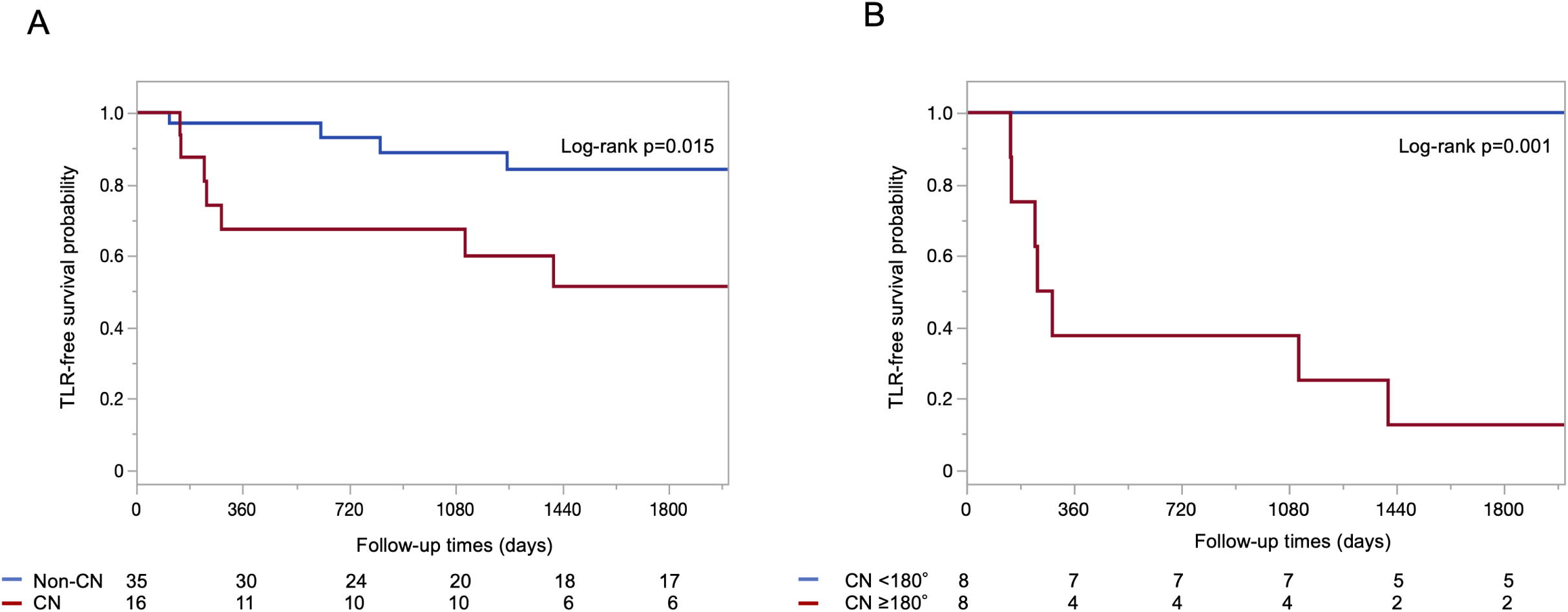
Kaplan–Meier curves of clinically-driven TLR. Kaplan–Meier curves showing the difference in the clinically-driven TLR-free survival between the CN and non-CN groups (A) and between CN angle ≥180° and <180° in the CN group. CN, calcified nodule; TLR, target lesion revascularization.

## DISCUSSION

In the present study, CN lesions were observed in 29.6% of the calcified lesions requiring RA. In-stent CNs were observed in 54% of the lesions treated for CN at the 8-month follow-up, where CN-like protrusion occurred, but were absent in non-CN lesions. Larger NIT at 8-month follow-up was associated with CN occurrence at index PCI but not with post-PCI stent expansion. Moreover, compared to patients with lesions without CNs, those with lesions with CNs treated with PCI using RA had a higher MACE incidence during the 5-year follow-up. Finally, the optimal cut-off value for the maximum CN arc to predict 5-year TLR was 182°.

Previous studies using intravascular ultrasound (IVUS) identified CN prevalence in 40.0–48.5% of calcified lesions requiring RA.^5,10,17^ In this study, we observed CNs in 29.6% of the target lesions using OCT. We defined CNs as eruptive CNs, excluding calcified protrusion (defined without eruptive nodules), as distinguishing between these two types of protruding Ca using IVUS can be challenging. Consistent with our results, several OCT studies demonstrated an association between CN occurrence and older age, renal dysfunction, and hemodialysis.^3,9^

In this study, in-stent CNs were present at the 8-month follow-up in 54% of the CN lesions at the initial PCI, whereas they were absent in the non-CN lesions. Consistent with our findings, Isodono et al. observed CN-like ISR findings in approximately 10% of all ISR lesions where severe calcification was evident on initial angiography before stent placement.^14^ Hamana et al. also identified in-stent CNs within an identical lesion where CNs were previously observed on OCT images.^9^ A previous pathological study identified two mechanisms for CN formation within the stent lumen: (1) CNs protruding from the underlying calcified plaque into the lumen and (2) *de novo* nodular calcification within the neointima protruding into the lumen.^7^ Unlike previous OCT studies, we observed target lesions with serial OCT assessments at pre-stent, post-PCI, and the 8-month follow-up. We confirmed that the in-stent CNs appeared where the CNs were located before stenting, and we observed CN-like protrusion after stenting, suggesting that CNs likely protruded through stent struts immediately after implantation. A previous OCT study also observed post-PCI protrusion of the CN in 58% of the eruptive CN lesions.^6^ These findings suggest that CNs protruding into the lumen from underlying calcified plaques may cause in-stent CNs.

In this study, CNs at the index PCI, rather than stent expansion, correlated with increased NIT, although stent under-expansion is the most consistent TLR predictor.^19,20^ PCI for CN lesions is associated with worse clinical outcomes than for non-CN lesions.^5,9,20^ In this study, the cumulative 5-year TLR incidence was 43.8% in CN lesions, higher than the previously reported values of 23.2%^5^ and 32.6%,^9^ possibly because of the differences in baseline Ca severity. In-stent CN contributes to TLR after PCI for CN lesions.^10,21^ In our study, the MLA site at follow-up corresponded to the CN site at baseline in 84.6% of the CN group, supporting the hypothesis that CN at index PCI was related to in-stent CNs and subsequent TLR. Moreover, TLRs are reported to predominantly occur within 1 year,^6,10^ and in-stent CNs are observed within the same timeframe after stenting.^8^ Consistently, in this study, TLR incidence in the CN group was predominantly observed within 1 year. Furthermore, persistent malapposition was frequently observed in the CN group, whereas most disappeared in the non-CN group, potentially contributing to increased TLR in CN lesions at long-term follow-up.^22^

OCT signal attenuation of CNs is associated with TLR at 5 years after stenting.^9^ In our study, among CN characteristics, the CN angle was associated with a 5-year TLR, and the optimal cut-off value for the maximum CN angle was 182°. Pathological studies indicated that calcified plaques invisible on OCT can harbor underlying CNs. Our study indicated that the CN angle, rather than length, may reflect the CN burden and is associated with increased TLR. Recent studies indicated that stentless PCI could be a potential therapeutic option for CNs when achieving an adequate luminal area without major dissection.^23–25^ Therefore, for CN lesions with maximum CN angles >182°, close follow-up and aggressive medical therapy may be required, and stentless PCI or bypass surgery may be a treatment option.

This study had several limitations. First, this was a retrospective observational study using a prospective single-center registry, and the study population was relatively small. Second, not all lesions could undergo preprocedural OCT owing to their severity. Third, selection bias might exist because 14 patients (25.9%) were excluded from the follow-up OCT analysis. Finally, the possibility of an occulostenotic reflex influenced by follow-up OCT findings cannot be ruled out. Further research is required to validate the importance and practicality of these findings in a broader population.

In conclusion, despite successful RA treatment with acceptable stent expansion, CN presence before PCI was associated with increased NIT formation with a new CN appearance, resulting in an increased TLR rate. The CN angle may be useful in stratifying stent failure risk.

### Source of Funding

None declared.

### Disclosures

M.O. declares lecture fees from Abbott Vascular, Asahi Intecc, Boston Scientific, and Terumo. The other authors declare no conflicts of interest.

### Supplemental Material

Detailed methods

Tables S1-S2

Figures S1–S3

## Non-standard Abbreviations and Acronyms

Ca: calcium
CN: calcified nodule
ISR: in-stent restenosis
IVUS: intravascular ultrasound
MACE: major adverse cardiac event
MLA: minimum lumen area
NIT: neointimal tissue
OCT: optical coherence tomography
PCI: percutaneous coronary intervention
RA: rotational atherectomy
TLR: target lesion revascularization

## Data Availability

Raw data were generated at Showa university Northern Yokohama Hospital. Derived data supporting the findings of this study are available from the corresponding author M.Y on request.

